## Supplementary figures and images for "Early Detection of Qualitative Fibrosis Progression Using Hybrid HU-Z-score CT Analysis: Overcoming Limitations of Traditional Quantitative Methods"

### SupplementaryFigure_S1_HU_Histograms.png

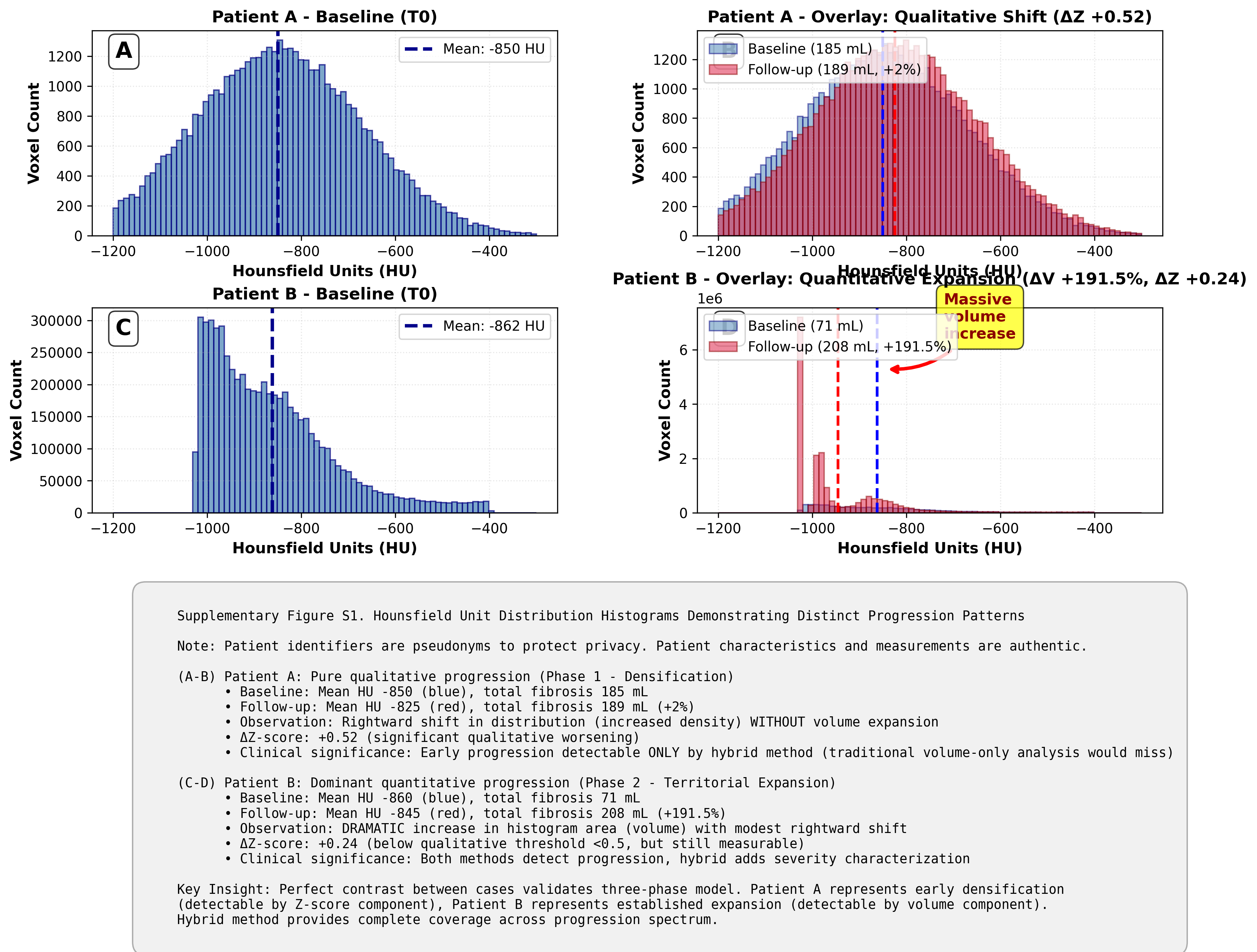
